# Blood cytokine analysis suggests that SARS-CoV-2 infection results in a sustained tumour promoting environment in cancer patients

**DOI:** 10.1101/2021.10.29.21265511

**Authors:** Fien HR De Winter, An Hotterbeekx, Manon Huizing, Angelina Konnova, Erik Fransen, Bart’s Jongers, Ravi Kumar Jairam, Vincent Van averbeke, Pieter Moons, Ella Roelant, Debbie Le Blon, Wim Vanden Berghe, Annelies Janssens, Willem Lybaert, Lieselot Croes, Christof Vulsteke, Surbhi Malhotra-Kumar, Herman Goossens, Zwi Berneman, Marc Peeters, Peter van Dam, Samir Kumar-Singh

## Abstract

Cytokines, chemokines and (angiogenic) growth factors (CCGs) have been shown to play an intricate role in the progression of both solid and haematological malignancies. Recent studies have shown that SARS-CoV-2 infection leads to worse outcome in cancer patients, especially in haematological malignancy patients. Here, we investigated how SARS-CoV-2 infection impacts the already altered CCG levels in solid or haematological malignancies, specifically whether there is a protective effect or rather a potentially higher risk for major COVID-19 complications in cancer patients due to elevated CCGs linked to cancer progression. Serially analysing immune responses with 55 CCGs in cancer patients under active treatment with or without SARS-CoV-2 infection, we first showed that cancer patients without SARS-CoV-2 infection (n=54) demonstrate elevated levels of 35 CCGs compared to the non-cancer, non-infected control group of health care workers (n=42). Of the 35 CCGs, 19 were common to both solid and haematological malignancy groups and comprised previously described cytokines such as IL-6, TNF-α, IL-1Ra, IL-17A, and VEGF, but also several less well described cytokines/chemokines such as Fractalkine, Tie-2, and T cell chemokine CTACK. Importantly, we show here that 7 CCGs are significantly altered in SARS-CoV-2 exposed cancer patients (n=52). Of these TNF-α, IFN-β, TSLP and sVCAM-1, identified to be elevated in haematological cancers, are also known tumour-promoting factors. Longitudinal analysis conducted over 3 months showed persistence of several tumour-promoting CCGs in SARS-CoV-2 exposed cancer patients. These data urge for increased vigilance for haematological malignancy patients as a part of long COVID follow-up.

## Introduction

Inflammation is one of the hallmarks of cancer. While chronic inflammation predisposes to development of specific cancers, cancer-related inflammation generally affects all aspects of malignancies, including proliferation and survival of malignant cells, angiogenesis, and metastasis [1-3]. Immune cell infiltration is observed in almost all tumours, ranging from subtle infiltrations detectable by only cell-type specific antibodies to gross accumulations of lymphocytes, macrophages, or mast cells [1-3]. These cell infiltrates secrete a repertoire of inflammatory proteins such as cytokines that serve as important orchestrators of cancer–inflammation interactions. For instance, cytokines like interleukin (IL)-2, IL-12, and certain interferons might inhibit the growth of tumours. However, paradoxically, many cytokines support chronic inflammation thereby promoting tumour growth and influence multiple aspects of cancer metastasis. Examples of such tumour promoting cytokines are IL-6, Tumour necrosis factor α (TNF-α) and Transforming growth factor β (TGF-β), where several clinical trials targeting these cancer-promoting cytokines are ongoing [3-5].

Several cytokines, chemokines, and growth factors (CCGs) are also currently being utilized in cancer patient care as prognostic classifiers [6-10]. A comprehensive CCG analysis is thus essential to untangle the individual interactions in the understanding of CCGs cumulative role on tumour growth and metastasis. While each cancer is associated with a distinct repertoire of cytokines, broad similarities might be present between solid and haematological malignancies, especially in the immunocompromised state of the metastatic phase [11,12]. For example, angiogenic factors are generally accepted to promote further growth of solid tumours but are also now increasingly found to be involved in haematological malignancies [13,14]. However, to our knowledge, a very limited number of studies have employed a comprehensive, quantitative analysis of CCGs in studies of cancer [7,9,15].

In the host defence system against infections, cytokines also play an important role in clearing viral or bacterial pathogens. Cancer patients are more susceptible to infections due to immunosuppression caused by cancer-mediated immune factors as well as systemic treatment. Thus, not surprisingly, cancer patients, especially those with advanced disease or haematological malignancies, were shown to be at higher risk of developing severe COVID-19 [16-18]. For instance, in the initial outbreak in Wuhan, a more severe disease was reported in hospitalized cancer patients than non-cancer patients, with higher rates of ICU admission, requirement for invasive ventilation, and higher mortality [16]. These data are in agreement with another study that, after correction for age and other co-morbidities, reported more severe COVID-19 in cancer patients, particularly in haematological and lung malignancies as well as in all metastatic stage IV cancers [17]. However, not all studies have confirmed these data [19] and it remains possible that the increased mortality in cancer patients is inherently biased by comorbidities such as old age, obesity and diabetes or inadequate therapy and follow up during the COVID-19 pandemic [20]. An equally relevant question is how CCGs are modulated in cancer patients after SARS-CoV-2 infection/exposure. Several pro-inflammatory cytokines observed to be massively upregulated in SARS-CoV-2 infection, such as IL-6 and TNF-α [21-25], are also tumour promoting factors and therefore could alter the rate of cancer progression. It is also possible that already elevated levels of cytokines such as IL-6 and TNF-α in cancer patients offer protection in the initial stages of SARS-CoV-2 infection, offering a certain degree of immune-preparedness. To the best of our knowledge, only a few studies have investigated this in the fragile cancer population [15].

Here, we characterised the immune response in cancer patients with solid and haematological malignancies exposed to SARS-CoV-2 compared to unexposed cancer patients matched for age-, gender- and cancer type, as well as unexposed health care workers (HCWs) from the same units that were matched for age category and gender. We aimed to investigate whether there is a protective effect or rather a higher risk for major COVID-19 complications in cancer patients. We also aimed to investigate baseline differences between solid and haematological malignancies in comparison with HCW controls without SARS-CoV-2 exposure.

## Materials and Methods

### Study design

Ambulatory cancer patients that were scheduled for routine blood sampling attending the Multidisciplinary Oncology Unit of Antwerp University Hospital (MOCA) between 24/3/2020 and 31/05/2020, and the Oncology Unit of AZ Maria Middelares Hospital, Ghent, between 13/04/2020 and 31/05/2020, and consenting to participate (n = 922), were studied prospectively as part of the first COVID-19 pandemic wave in Belgium [26]. In addition, health care workers (HCWs; n = 92) from these units donated similar blood samples during the study period at time points 0, 1, 2 and 3 months after written informed consent. Active SARS-CoV-2 infection was diagnosed by detection of viral RNA in nose/throat swabs by the Cobas SARS-CoV-2 RT-PCR (Roche) on the automated Cobas 6800 system. Anti-SARS-CoV-2 immunoglobins (Ig) in blood were tested by three commercial tests, namely, Liaison SARS-CoV-2 S1/S2 IgG (DiaSorin, Saluggia, Italy), Alinity SARS-CoV-2 IgG (Abbott, Chicago, IL, USA), and Elecsys Anti-SARS-CoV-2 (Roche, Basel, Switzerland), as described [26].

A total of 44 cancer patients showed evidence of SARS-CoV-2 exposure with either PCR or serology and were enrolled in the current CCG study. In addition, SARS-CoV-2 PCR-positive ambulatory cancer patients attending the Multidisciplinary Oncology Unit between 1/6/2020 and 31/12/2020 as part the second COVID-19 wave were included (n = 10). Lastly, from a third Belgian Oncology Unit at the AZ Nikolaas, Sint-Niklaas, additional SARS-CoV-2 exposed cancer (n = 8) patients were sampled after written informed consent between 5/11/2020 and 1/12/2020, also as part of the second COVID-19 wave. From these 62 cancer patients, 10 could not be studied as either no post-COVID-19 sample was available (n = 6) or the patients died due to COVID-19 consequences without a blood sample for analysis (n = 4). Thus, a total of 52 cancer patients were analysed for CCGs in this study. Similarly, from 92 HCW enrolled in the first wave, 19 (20.7%) were SARS-CoV-2 positive in the first or second wave and were enrolled in the study. For 15 HCWs, a blood sample was available after SARS-CoV-2 exposure, and these were studied for blood CCGs. Besides this, unexposed cancer patients matched for age, gender and cancer type (n = 54) and unexposed HCW matched for age, gender and co-morbidity (n = 42) were selected as controls.

Clinical data including SARS-CoV-2 related symptoms were recorded upon enrolment in the study and peak disease severity was determined according to the World Health Organization COVID-19 ordinal scale [27]. The study was approved by the ethics committee of the Antwerp University hospital (EC number 20/13/156, internal EDGE 001070).

### Sample processing

For CCG immunoassays, whole blood was prospectively collected in 10 mL EDTA tubes (BD Vacutainer K2E), and plasma was prepared. Briefly, within 3 hours of blood collection, samples were centrifuged at 1900 g for 10 minutes without brakes. After the first spin, plasma was transferred to a new tube and again spun at 1900 g for 10 minutes without brakes. Aliquots were flash frozen in liquid nitrogen and stored in the Biobank of Antwerp University Hospital at −80°C until the multiplex analysis.

### CCG measurements in plasma

CCGs were measured in EDTA plasma samples using U-plex and V-plex panels from Meso Scale Discovery (MSD, MD, USA) according to the manufacturer instructions. The following 55 CCGs were measured: Brain-derived neurotrophic factor (BDNF), Basic fibroblast growth factor (bFGF), C-reactive protein (CRP), Cutaneous T-cell attracting chemokine (CTACK), Eotaxin, Erythropoietin (EPO), Vascular endothelial growth factor receptor 1 (FlT-1), Fractalkine, Granulocyte colony stimulating factor (G-CSF), Granulocyte-macrophage colony-stimulating factor (GM-CSF), Macrophage colony-stimulating factor (M-CSF), Interferon β (IFN-β), Interferon γ (IFN-γ), Interleukin (IL)-1β, IL-1 receptor antagonist (IL-1Ra), IL-2, IL-2 receptor α (IL-2Rα), IL-4, IL-5, IL-6, IL-7, IL-8, IL-9, IL-10, IL-12p40, IL-12p70, IL-13, IL-15, IL-16, IL-17A, IL-17F, IL-18, IL-21, IL-22, IL-23, IL-33, IFN-γ induced protein 10 (IP-10), Monocyte chemoattractant protein (MCP)-1, MCP-2, MCP-3, Macrophage inflammatory protein (MIP)-1α, MIP-1β, MIP-3α, Placental growth factor (PlGF), Serum amyloid A (SAA), soluble intercellular adhesion molecule 1 (sICAM-1), soluble vascular cell adhesion molecule 1 (sVCAM-1), active and total (acid activated) tumour growth factor β (TGF-β), Angiopoietin receptor 1 (Tie-2), Tumour necrosis factor α (TNF-α), Thymic stromal lymphopoietin (TSLP), Vascular endothelial growth factor (VEGF)-A, VEGF-C and VEGF-D.

Measurements were performed in randomized batches. Briefly, 96-well plates of the U-plex panels were coated with a capturing antibody linked to a linker for one hour. The vascular injury panel (K15198D) was washed before use. The angiogenesis panel (K15190D) was blocked with MSD blocking buffer A for one hour. All plates were then washed three times with PBS-Tween (0.05%). Samples were incubated for one hour (except for the angiogenesis and the vascular injury panels, as well as the BDNF panel where two hours of incubation were performed), after which the plates were washed three times again. Detection antibody with a sulfo-tag was added and after another one-hour incubation step (two hours for the angiogenesis panel) plates were washed and read with MSD reading buffer on the QuickPlex SQ 120 (MSD).

### Statistics

All data were analysed using SPSS v27, R (version R4.0.4) and Metaboanalyst 5.0 (https://www.metabo-analyst.ca/). CCG levels were measured in plasma samples at different timepoints for most individuals. For comparison between groups, the sample timepoint closest to the diagnosis of SARS-CoV-2 was used for the exposed individuals and the average of all samples was used for the unexposed individuals. Patient and HCW parameters were compared by the non-parametric Kruskal-Wallis test for continuous variables and Fisher’s exact test for categorical variables. Group differences in CCG profiles were explored by Partial Least-Squares Discriminant Analysis (PLS-DA) using Metaboanalyst. This analysis was performed on data normalised by autoscaling and natural log transformation.

HCWs without SARS-CoV-2 exposure were compared with unexposed oncology patients grouped according to type of haematological and solid malignancies. One-way Analysis of variance (ANOVA) was carried out to test for differences in the CCG levels between different groups followed by a post-hoc analysis with Tukey correction for multiple hypothesis testing. The test was carried out on log10-transformed values due to the non-normality of the outcome data. Reported effect sizes and fold changes were backtransformed. The significance of the test was assessed using a False discovery rate (FDR) analysis to account for the multitude of hypotheses tested, with q-values indicating the fraction of false positive associations if a given p-value is declared significant.

To model the change in CCG concentrations over time after SARS-CoV-2 exposure in the three groups, all timepoints after exposure were included in a linear mixed model with the logarithm of CCG concentrations as dependent variable, time as fixed effect and individual identity as random effect to account for the non-independence of observations within the same individual. To investigate whether there was a different evolution of the CCG concentration over time between the groups, linear mixed models were fitted with time, cancer type and their interaction as fixed effect, with the p-value of the interaction testing the null hypothesis that the slope was the same across all groups. In case the slope was significantly different between the groups (p-value for interaction < 0.05), we carried out a pairwise comparison of the slopes between the HCWs and solid tumours and between HCWs and haematological malignancies. Starting from the slope estimates and their standard error, obtained through a linear mixed model of CCG versus time, a Z-test was carried out testing the null hypothesis that the slope in the two tested groups were the same. Since two pairwise comparisons were made, a Bonferroni-correction was carried out on the resulting p-values.

## Results

### Patient characteristics

A total of 52 cancer patients positive for SARS-CoV-2 PCR or serology from the first and second COVID-19 waves in Belgium were studied. Of these, 36 (69.2%) patients had a solid tumour and 16 (30.8%) had a haematological malignancy (**Table 1 and SI Table 1**). Similarly, 15 HCWs who were SARS-CoV-2 positive in this study period were included in this study. Unexposed cancer patients (n = 54) who were matched with exposed cancer patients for age, gender and cancer type were also enrolled (**SI Table 2**). Of all cancer patients, 68 patients had solid tumours and 38 haematological malignancies. Lymphomas were the most common malignancy (n = 17, 16%), followed by breast cancer (n = 16, 15%), gastric cancers (n = 10, 9.4%), head and neck cancers (n = 9, 8.5%), leukaemias (n = 8, 7.5%) and respiratory tract tumours (n = 6, 5.7%). All other malignancy types were present in less than 5% of the patients (SI Table 2 for overview of cancer types). As healthy controls, 42 unexposed HCWs were enrolled that were group-matched with exposed HCWs for co-morbidity, age, and gender.

**Table 1.** Patient characteristics*

|  | Solid cancer (n = 68) |  | Haematological cancer (n = 38) |  | HCWs (n = 57) |  |
| --- | --- | --- | --- | --- | --- | --- |
|  | n (%) or mean (SD) |  | n (%) or mean (SD) |  | n (%) or mean (SD) |  |
|  | Exposed<sup>a</sup> | Unexp<sup>b</sup> | Exposed<sup>a</sup> | Unexp<sup>b</sup> | Exposed<sup>a</sup> | Unexp<sup>b</sup> |
| SARS-CoV-2 status | n = 36 | n = 32 | n = 16 | n = 22 | n = 15 | n = 42 |
| Asymptomatic | 8 (22.2%) | NA | 5 (31.3%) | NA | 6 (40.0%) | NA |
| Mild | 19 (52.8%) | NA | 6 (37.5%) | NA | 7 (46.7%) | NA |
| Moderate | 3 (8.3%) | NA | 2 (12.5%) | NA | 0 (0%) | NA |
| Severe | 6 (16.7%) | NA | 2 (12.5%) | NA | 2 (13.3%) | NA |
| Critical | 0 (0%) | NA | 1 (6.3%) | NA | 0 (0%) | NA |
| Male sex | 15 (41.7%) | 17 (53.1%) | 7 (43.8%) | 13 (40.6%) | 2 (13.3%) | 5 (11.9%) |
| Age, mean (SD) | 58.8 (13.6) | 60.5 (12.7) | 57.9 (13.9) | 53.6 (20.4) | 37.5 (11.4) | 35.5 (10.7) |
| BMI, mean (SD) | 25.7 (6.1) | 25.6 (6.4) | 24.6 (6.9) | 24.1 (4.0) | 22.9 (0.0) | 24.2 (3.7) |
| Recent chemotherapy | 22 (61.1%) | 20 (62.5%) | 11 (68.8%) | 15 (46.9%) | 0 (0%) | 0 (0%) |
| Radiotherapy | 8 (22.2%) | 5 (15.6%) | 3 (18.8%) | 3 (9.4%) | 0 (0%) | 0 (0%) |
| Recent targeted therapy | 12 (33.3%) | 9 (28.1%) | 8 (50%) | 9 (28.1%) | 0 (0%) | 0 (0%) |
| Antihormonal treatment | 2 (5.6%) | 3 (9.4%) | 1 (6.3%) | 0 (0%) | 0 (0%) | 0 (0%) |
| Transplant | 0 (0%) | 0 (0%) | 4 (25.0%) | 5 (15.6%) | 0 (0%) | 0 (0%) |

HCWs: Health care workers
SD: standard deviation
<sup>a</sup>All SARS-CoV-2 exposed individuals were included for CCG analysis
<sup>b</sup>Unexposed individuals were group-matched to exposed individuals based on age, gender and cancer type in case of cancer patients
NA: not applicable
\*Additional patient comorbidities are represented in Supplementary Table 1

No significant differences were observed in patients with solid or haematological malignancies for sex and age (Table 1). When comparing both cancer types with HCWs, there were significantly fewer males in the HCW cohorts (12.3% males) compared to both cancer groups (solid unexposed/exposed 53.1%/41.7%; haematological unexposed/exposed 40.6%/43.8%). Additionally, HCWs were significantly younger (Average (Av) 36 years, *P* < 0.001) than both groups of cancer patients (solid cancer Av 59.6 years; haematological malignancy Av 55.4 years). BMI was not significantly different between groups (solid cancers Av 25.6; haematological malignancies Av 24.3; HCWs Av 24.1). None of the HCWs were diabetic compared to 8 (11.8%) in the solid and 1 (2.6%) in the haematological malignancy patient groups. Furthermore, there were fewer infections other than SARS-CoV-2 in HCWs (3.5% overall) compared to solid tumour patients (16.2% overall) and haematological malignancy patients (28.9% overall) (SI Table 1). Nine patients in the haematological malignancy group received bone marrow transplantation (n = 9). Cancer patients received chemotherapy (n = 67), antihormonal therapy (n = 6) or targeted therapy such as monoclonal antibodies, proteasome inhibitors, signal transduction inhibitors and angiogenesis inhibitors (n = 37).

### No significant difference in disease severity between SARS-CoV-2 exposed cancer patients and exposed health care workers

We studied whether SARS-CoV-2 exposed cancer patients or HCWs in our cohorts had a different disease severity. Disease severity for the patients enrolled for the CCG study was graded as asymptomatic, mild, moderate, severe, and critical [27](Table 1). For this analysis, we also included the 10 additional patients that were excluded for the CCG study for the lack of plasma samples. While no significant difference was observed for any of the severity groups between HCWs and the cancer groups, 89.5% of HCWs and 71.1% and 82.4% of the solid and haematological malignancy group, respectively, remained asymptomatic or showed mild to moderate symptoms **(SI Figure 1**, Table 1). Although a slightly higher disease severity is observed in the solid cancer cohort, this difference was not significant (Fisher’s exact test). Bearing in mind that cancer patients were approximately 20 years older, and gender balanced (males 49.1%) while HCWs were mostly females (87.7%), these data suggest that cancer patients are not significantly different for disease severity in the cohort studied here.

### In absence of SARS-CoV-2 exposure, CCG profiles in solid and haematological malignancies are not inherently different from each other, but different from unexposed healthy controls

We analysed plasma samples of unexposed cancer patients with either solid or haematological malignancies as well as unexposed HCWs for 55 CCGs (**Figure 1**). Partial least squares discriminant analysis (PLS-DA) identified a clear clustering of the HCWs and combined group of cancer patients (Accuracy = 91%, R^2^ = 0.76, Q^2^ = 0.63 for 2 components; Figure 1A). High classification accuracies were also achieved between the unexposed HCW group and unexposed solid tumours (Accuracy 86%, R^2^=0.71, Q^2^=0.58; Figure 1B) and between the unexposed HCWs and haematological malignancies (Accuracy 91%, R^2^=0.76, Q^2^=0.63; Figure 1C). However, we did not find a clear clustering of the CCG profiles between haematological and solid malignancies (Q^2^= −0.33 for 2 components; Figure 1D). Further analysing the individual cytokine differences between solid and haematological malignancies with a one-way analysis of variance (ANOVA), we found a statistically significant increase in only 3 CCGs in patients with solid tumours compared to those with haematological malignancies: BDNF (4.1-fold, *P* = 0.004), VEGF-C (2.2-fold, *P* = 0.019), and SAA (2.2-fold, *P* = 0.016). These data suggest that while the baseline CCG profiles in unexposed patients with solid or haematological malignancies are significantly different from the unexposed HCWs, CCG profiles of unexposed patients with either solid or haematological malignancies are not grossly distinct.

**Figure 1.**
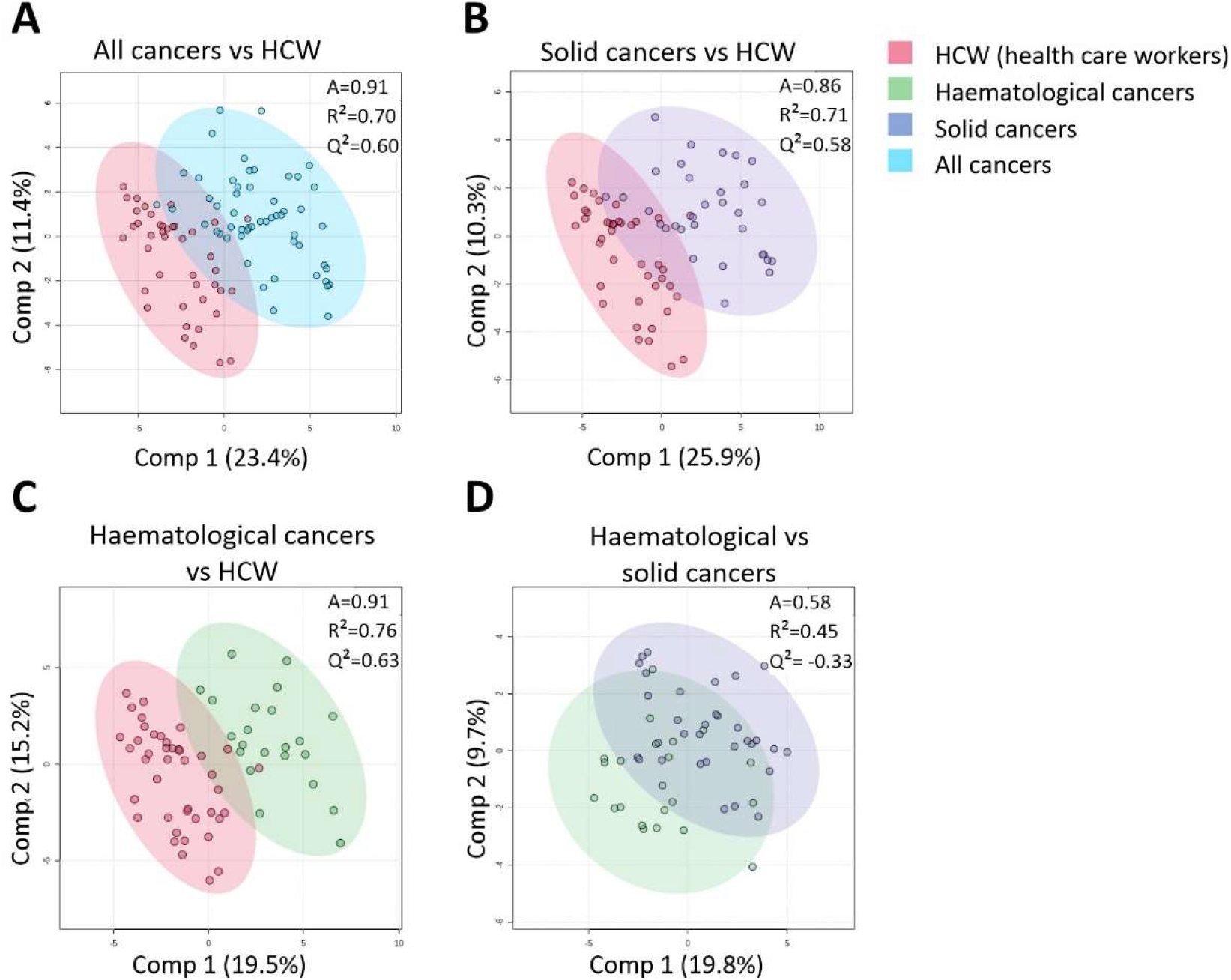
Cluster analysis of CCGs in unexposed individuals reveals larger differences between health care workers (HCWs) and cancer as a group or individually as solid and haematologic groups, than between the two cancer groups. A) The comparison of HCWs with the two different cancer types pooled. B) The comparison of the HCWs with patients with solid tumours C) and with haematological malignancies D) Comparison of haematological versus solid cancer patients. A, accuracy.

### A broad group of inflammatory markers and growth factors distinguishes unexposed solid or haematological malignancies from healthy controls

Next, we studied the basis of the discriminant CCG profiles between both groups of unexposed cancer patients and unexposed HCWs by ANOVA. These results are presented in **Figures 2–5, SI Figure 2 and 3A, and SI Table 3A**. First, consistent with the role of inflammation in cancer development and progression [2], we observed a significant upregulation of Th1 pro-inflammatory cytokines in both solid and haematological malignancy patients compared to HCWs, including TNF-α, IL-6, and IL-1Ra (**Figure 2A**). A significant upregulation of neutrophilic chemokine IL-8 and acute phase proteins CRP and SAA was also observed for solid tumours, but not for haematological malignancy patients (**Figure 2B**). Regarding the interferon family, while IFN-β and IP-10 were not significantly altered, IFN-γ was significantly increased in both solid and haematological malignancy patients, and IL-18 (also called IFN-γ-inducing factor) only in the later cancer group (**Figure 2C**). Similarly, pro-inflammatory Th17-related cytokines, IL-17A and IL-22 were significantly upregulated in both solid and haematological malignancy patients compared to HCWs (**Figure 2D**).

**Figure 2.**
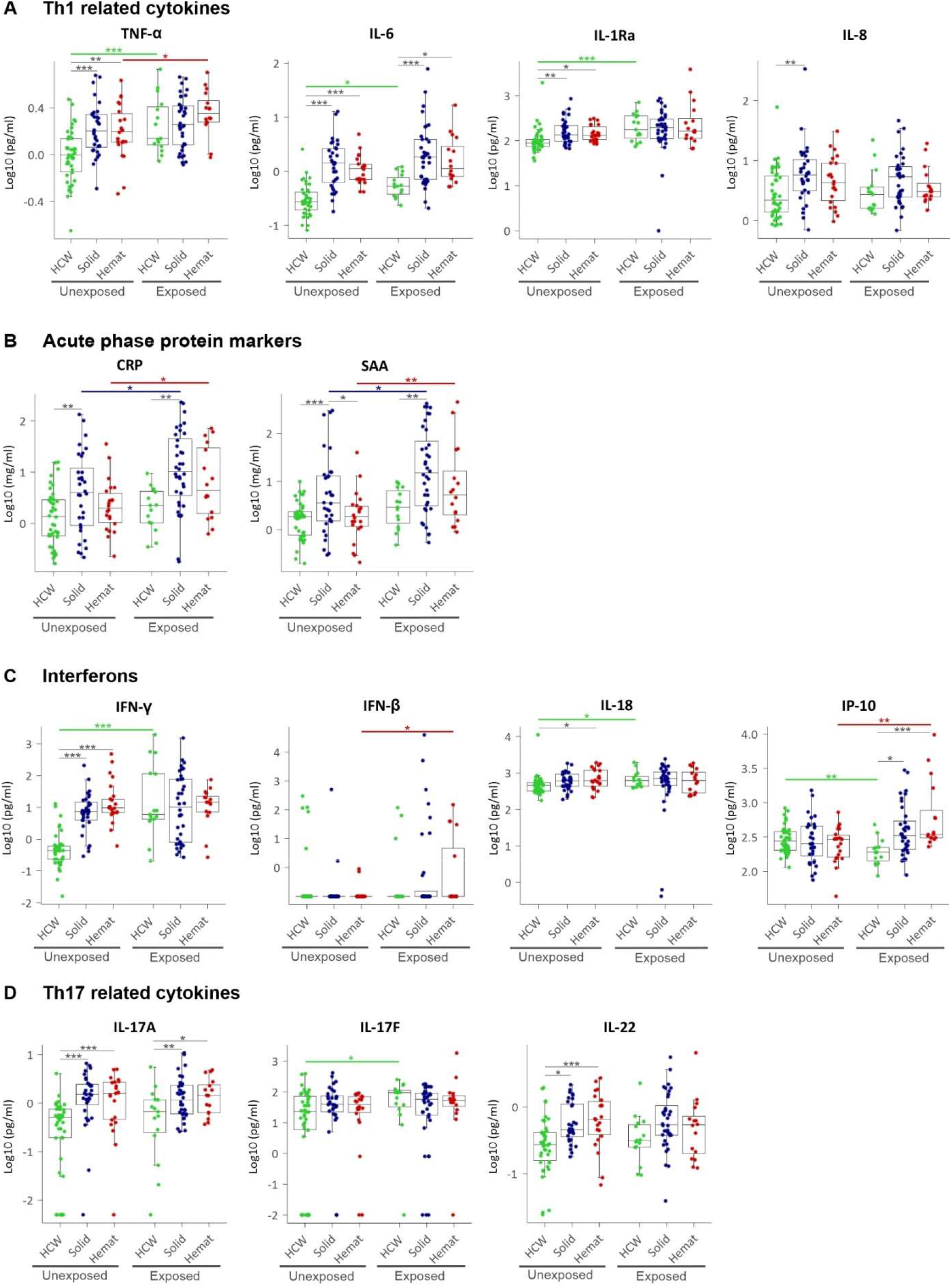
Plasma levels of pro-inflammatory CCGs as A) Th1 related cytokines, B) Acute phase proteins, C) Interferons and interferon-related cytokines, and D) Th17 related cytokines. HCW, health care workers (green), Solid, patients with solid tumours (blue), Hemat, patients with haematological malignancies (red). Each dot represents the sample closest to exposure in case of exposed patients, and an average of multiple timepoints, when available, for unexposed patients. * *P* < 0.05, ** *P* < 0.01, *** *P* < 0.001.

We further studied the Th2 family of cytokines which besides being anti-inflammatory are promotors of tumour cell growth and invasion by enhancing pro-tumour properties of macrophages [28]. We observed a significant upregulation of IL-33, TSLP and IL-9 for both solid and haematological malignancy patients compared to HCWs, and for IL-5, eotaxin and IL-21 in only solid tumour patients (**Figure 3A**). However, no significant differences in the major Th2 cytokine IL-4 and IL-13 levels were observed for either of the cancer groups compared to HCWs. For immunomodulatory cytokines, IL-10 and IL-15 were found to be significantly upregulated in both solid and haematological malignancy patients compared to HCWs, while another Treg associated cytokine, IL-2Rα, was significantly upregulated only in solid tumour patients (**Figure 3B**).

**Figure 3.**
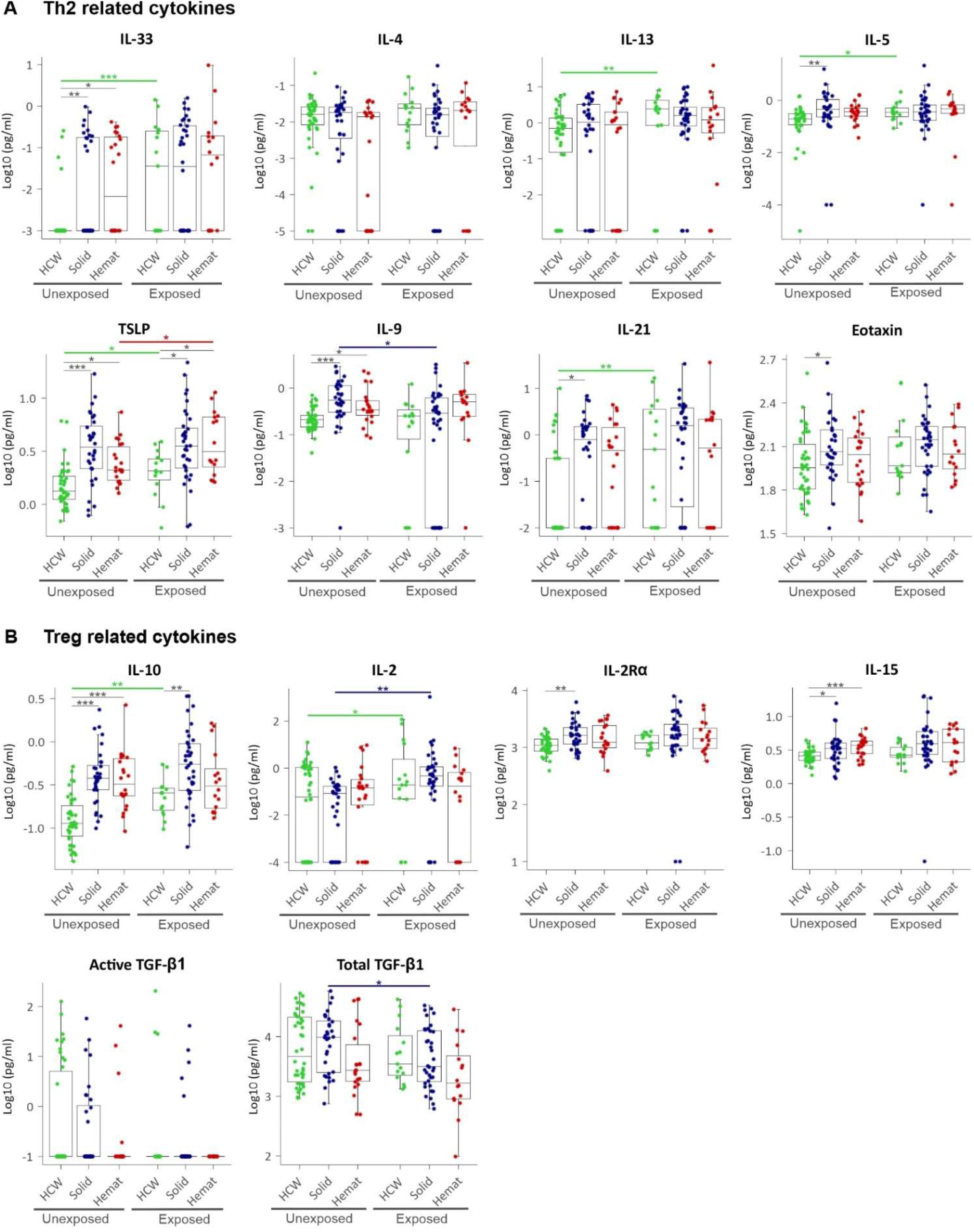
Plasma levels of A) Th2 related (anti-inflammatory) cytokines and B) Treg-related (immunomodulatory) cytokines. HCW, health care workers (green), Solid, patients with solid tumours (blue), Hemat, patients with haematological malignancies (red). Each dot represents the sample closest to exposure in case of exposed patients, and an average of multiple timepoints, when available, for unexposed patients. * *P* < 0.05, ** *P* < 0.01, *** *P* < 0.001.

We also addressed angiogenic growth factors (i.e., VEGF family, bFGF, PlGF), their receptors (Flt-1, Tie-2), haematopoietic stimulators (EPO, IL-7), as well as adhesion molecules expressed by endothelial cells (sICAM, sVCAM). Besides this, we also studied BDNF, a newly identified mediator of angiogenesis that acts through stimulating VEGF [29]. While angiogenesis plays a critical role in progression of solid tumours, it is increasingly recognized that haematological malignancies also depend on the induction of new blood vessel formation [14]. Consistent to these data, we showed that while VEGF-A and its receptor Flt-1, bFGF and ICAM were significantly upregulated for solid tumours, PlGF and EPO were significantly upregulated for both solid and haematological malignancy patients compared to HCWs. Interestingly, angiopoietin receptor Tie-2 was significantly 30-35% downregulated in both solid and haematological malignancy patients compared to HCWs (**Figure 4**).

**Figure 4.**
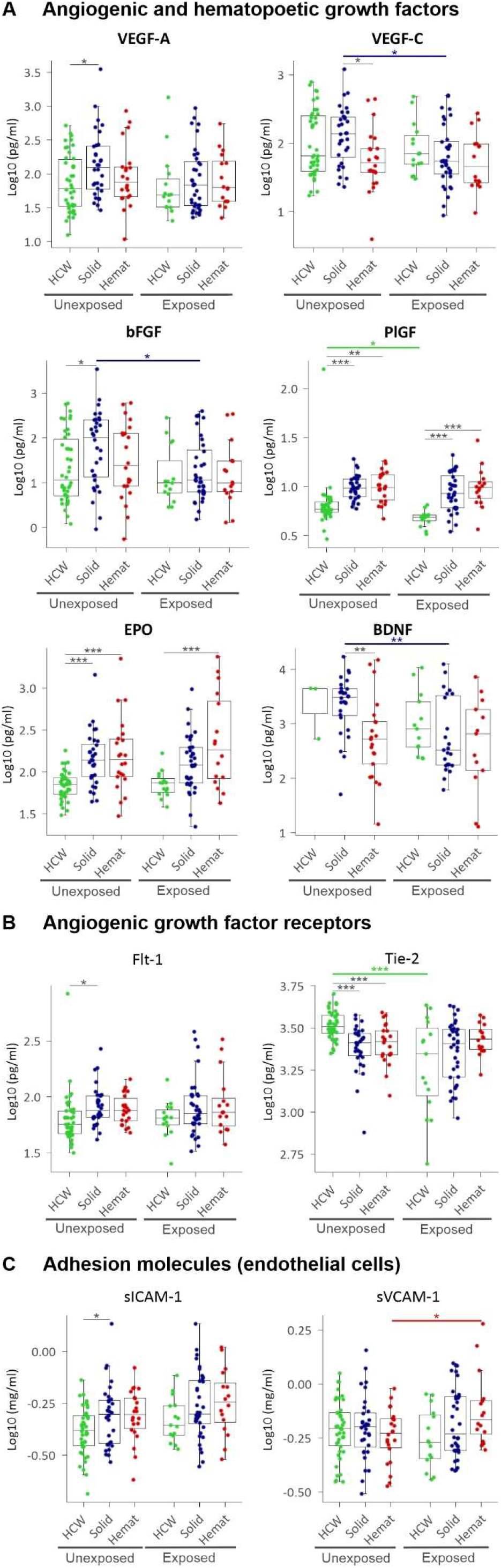
Plasma levels of A) Angiogenic and haematopoietic growth factors, B) Angiogenic growth factor receptors, and C) Adhesion molecules expressed by endothelial cells. HCW, health care workers (green), Solid, patients with solid tumours (blue), Hemat, patients with haematological malignancies (red). Each dot represents the sample closest to exposure in case of exposed patients, and an average of multiple timepoints, when available, for unexposed patients. * *P* < 0.05, ** *P* < 0.01, *** *P* < 0.001.

Lastly, we addressed the question whether colony stimulating factors and chemokines were distinctly altered in solid or haematological malignancies. We first showed that colony stimulating factors for neutrophils (G-CSF) and macrophages (M-CSF) were significantly upregulated in both solid and haematological malignancy patients, compared to HCW controls, while GM-CSF, a broader stimulant for all granulocytes and monocytes, was only upregulated in solid cancer patients (**Figure 5A)**. Concerning chemokines, we showed that monocyte chemotactic proteins MCP-1 and −2—considered as principal chemokines involved in the recruitment of monocytes/macrophages and activated lymphocytes—were significantly upregulated in both solid and haematological malignancies, while closely related MCP-3 and MIP-1β /CCL4 implicated in the chemotaxis of dendritic cells and eosinophils were only upregulated in solid tumours but not haematological malignancies. Similarly, CCL20/MIP-3α a chemotactic factor for lymphocytes was also upregulated only in solid tumours, while CCL27/CTACK was upregulated for both solid and haematological malignancies. Lastly, CX3CL1/fractalkine, a chemokine abundantly expressed by activated endothelium and promoting strong adhesion of leukocytes to endothelial cells which is also involved in thrombosis was significantly upregulated by up to 30% in both solid and haematological malignancy patients compared to HCWs (**Figure 5B**). These data suggest gross alterations in CCG profiles of solid or haematological malignancy patients, with one important mediator, Tie-2, remarkably downregulated.

**Figure 5.**
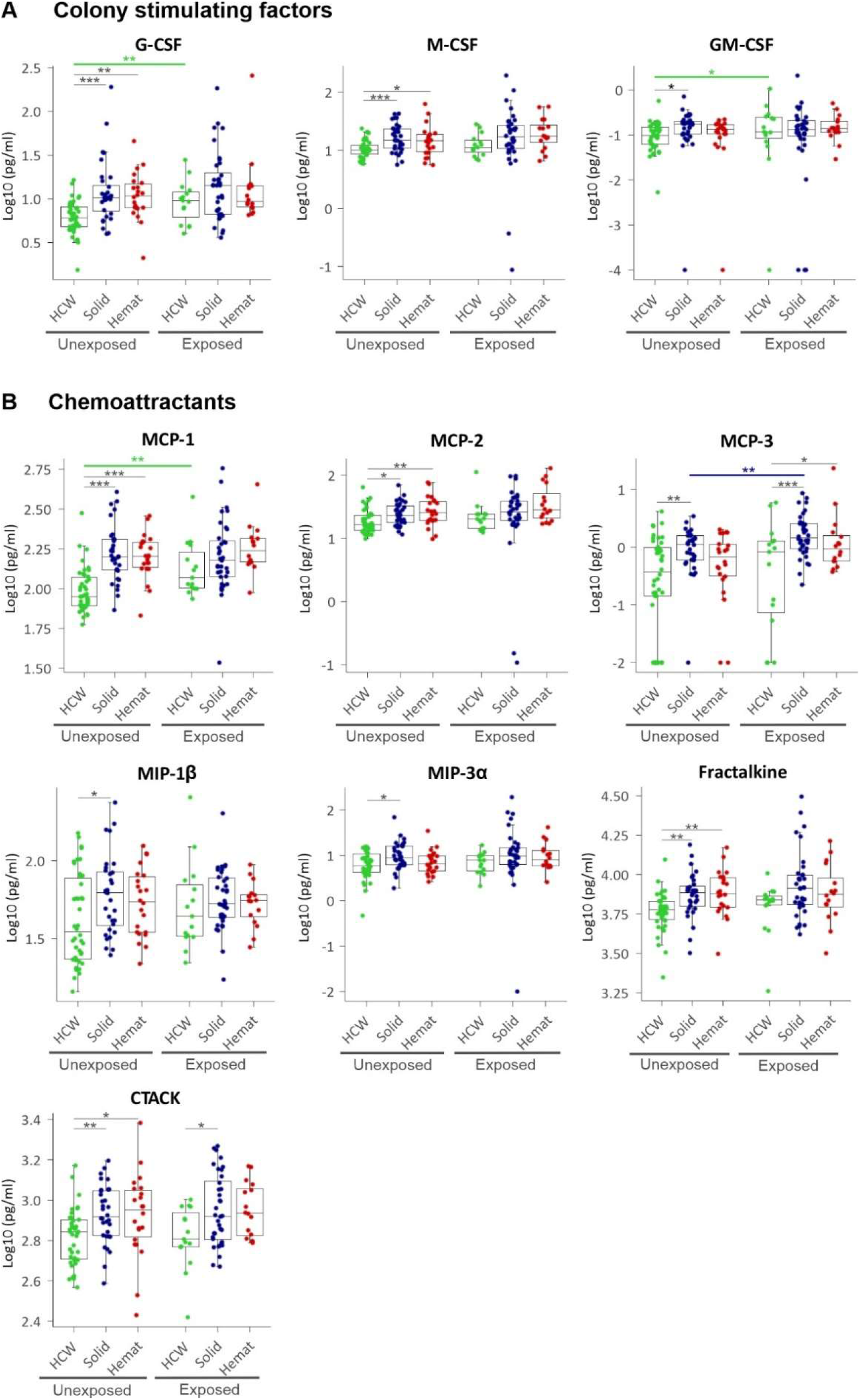
Plasma levels of A) Colony stimulating factors (CSFs) and B) Monocyte chemoattracting proteins (MCPs), macrophage inflammatory proteins (MIPs) and other chemokines. HCW, health care workers (green), Solid, patients with solid tumours (blue), Hemat, patients with haematological malignancies (red). Each dot represents the sample closest to exposure in case of exposed patients, and an average of multiple timepoints, when available, for unexposed patients. * *P* < 0.05, ** *P* < 0.01, *** *P* < 0.001.

### SARS-CoV-2 exposure elicits an expected increase in CCG levels in HCWs

Next, we studied how CCG levels in plasma of SARS-CoV-2 altered in exposed HCWs compared to those who were unexposed. Here, significantly higher levels of the Th1 cytokines and acute phase proteins—TNF-α (1.4-fold), IL-1Ra (2.0-fold), IL-6 (1.2-fold) and SAA (1.5-fold)—were observed in exposed HCWs compared to unexposed HCWs (**Figure 2A, B; SI Figure 3B**, SI Table 4A). For the interferon family, IFN-γ was significantly increased (11.8-fold) whereas IP-10 was significantly decreased (1.5-fold) (**Figure 2C**) along with the proinflammatory IL-17F (3-fold increased) (**Figure 2D**). Interestingly, most Th2 cytokines were also significantly increased in HCWs after exposure to SARS-CoV-2 and included IL-33 (1.2-fold), IL-5 (1.2-fold), IL-13 (1.6-fold), TSLP (1.2-fold) and IL-21 (1.8-fold) (**Figure 3A**). Immunomodulatory IL-10 was also found to be slightly but significantly increased in exposed HCWs (1.1-fold) along with Treg-associated IL-2 (1.9-fold; **Figure 3B**). While exposed HCWs showed a decrease in angiogenic growth factor PlGF (1.3-fold) and angiopoietin receptor Tie-2 (1.7-fold) (**Figure 4**), an increase was found for G-CSF (1.4-fold), GM-CSF (1.1-fold) and monocytes/macrophages chemoattractant MCP-1 (1.4-fold) (**Figure 5**). Although we predominantly studied post-acute timepoints and the patients were mostly in milder categories, these data are in agreement with COVID-19 where similar alterations in CCGs are observed in severe COVID-19 at acute timepoints [21-25,30-33].

### SARS-CoV-2 exposure causes noteworthy alterations in CCG profiles of solid and haematological malignancy patients

More central to the main hypothesis of the paper, we further studied whether SARS-CoV-2 exposure in cancer patients elevates CCGs linked to cancer progression. Despite elevated levels of several inflammatory mediators already occurring in cancer patients in comparison to non-cancer controls, 7 CCGs were additionally found to be significantly altered in cancer patients exposed to SARS-CoV-2. Specifically, in patients with solid tumours, besides a significant upregulation of the inflammatory markers CRP (2.1-fold) and SAA (2.5-fold), only immune cell activators IL-2 (1.9-fold) and MCP-3 (1.3-fold) were elevated (**Figure 2–5**, SI Figure 3B, SI Table 4B). In contrast, 5 CCGs showed a significant reduction in solid malignancy patients exposed to SARS-CoV-2, namely, angiogenesis growth factors VEGF-C (1.9-fold), bFGF (2.9-fold), and BDNF (3.7-fold), as well as IL-9 (1.2-fold) and total TGF-β (1.8-fold).

Similar to patients with solid tumours, individuals with haematological tumours exposed to SARS-CoV-2 showed a significant increase in the inflammatory markers CRP (2.4-fold) and SAA (3.1-fold). Furthermore, there was an increase in TNF-α (1.3-fold), IP-10 (2.5-fold), TSLP (1.4-fold), VCAM-1 (1.1-fold) and antiviral IFN-β (2.7-fold) (**Figure 2–5**, SI Table 4C).

### Longitudinal analysis shows persistence of CCG alterations in SARS-CoV-2 exposed cancer patients

Because several CCGs are involved in tumour progression, we studied the temporal evolution of the 14 CCGs for which we showed a significant increase or decrease for the exposed cancer groups. Samples from multiple timepoints collected over three months were available for the exposed cancer and HCW groups and were analysed. CCG levels were log transformed and entered in a linear mixed model to test whether the 14 CCGs for the cancer and HCW groups significantly changed over time and whether the rate of change for solid cancer or haematological malignancy groups significantly differed from that of the HCW group.

We first showed that while the inflammatory mediators CRP and SAA significantly declined in HCWs after SARS-CoV-2 exposure, no significant decline was observed for the exposed solid or haematological malignancy groups suggesting a persistence of pro-inflammatory state in cancer patients exposed to SARS-CoV-2 (**Figure 6**). A persistence of CCGs such as TNF-α, IL-2, and MCP-3 was also observed for exposed cancer patients but not for the healthy control group. For the haematological malignancy patient group, levels of IL-2 and MCP-3 even temporarily increased in the three-month follow-up and was significantly different from the declining levels noted for healthy controls. A significant temporal increase in IP-10 was also observed for haematological malignancy patients compared to a non-significant declining trend for healthy controls. These data suggest that several CCGs shown to be significantly increased in the exposed cancer patients compared to unexposed cancer patients are in fact long-lasting changes that could persist for at least 3 months after SARS-CoV-2 exposure.

**Figure 6.**
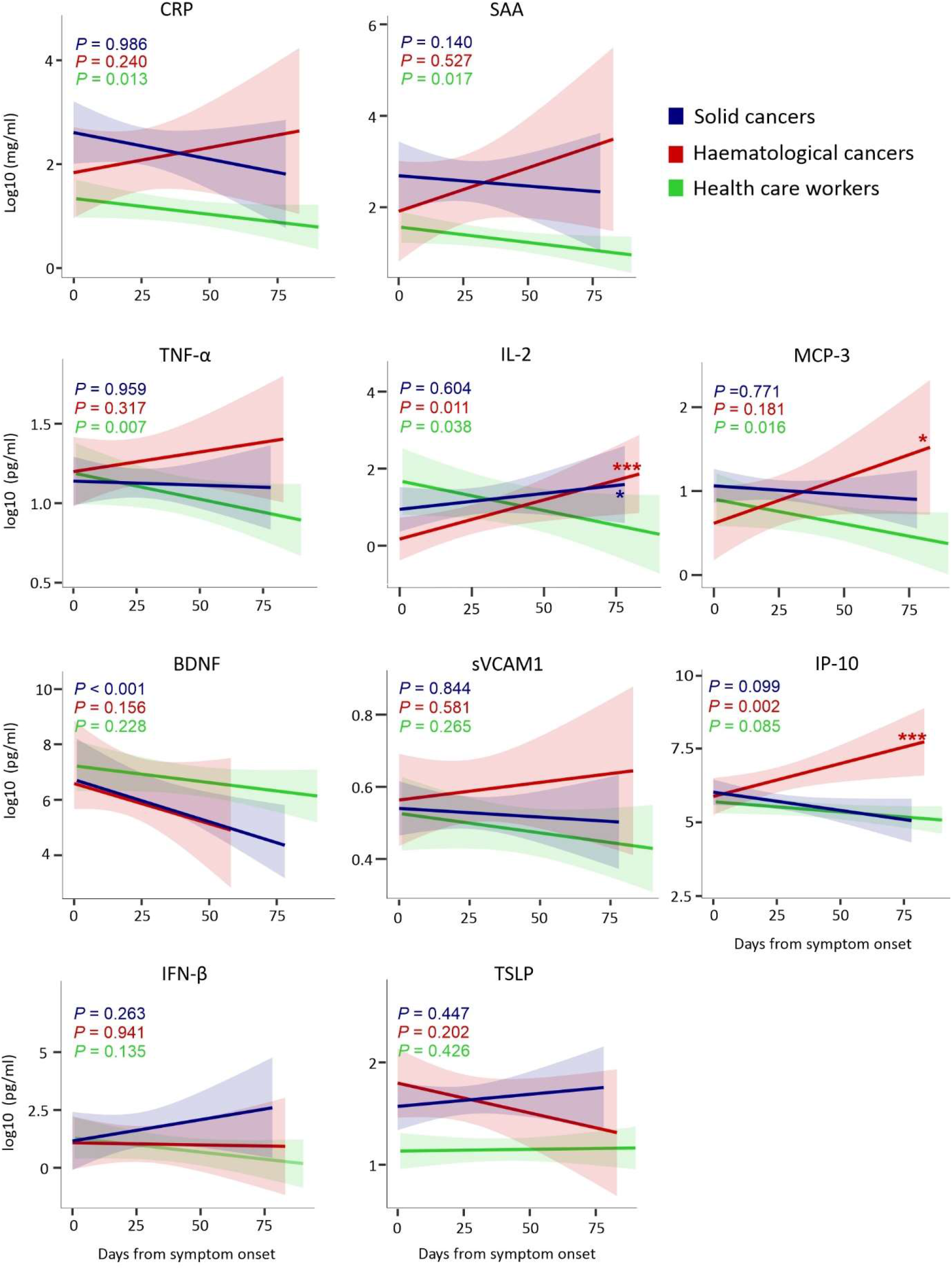
Temporal evolution of CCGs altered in exposed cancer patients. HCW, health care workers (green), Solid, patients with solid tumours (blue), Hemat, patients with haematological malignancies (red). Time is represented as days since symptom onset (day 0). *P*-values in the graph refer to significance of the slope from the 3 separate regression lines. The asterisks represent the significance of the pairwise comparison between the slope (p-value for interaction) in haematological malignancy versus HCW (red) and solid malignancy versus HCW (blue). \**P* < 0.05, ** *P* < 0.01, *** *P* < 0.001

## Discussion

In a highly diverse set of cancer types representing the most common types of solid as well as haematological malignancies, we identified distinct immunological profiles of CCGs in these two types of cancers. While 19 CCGs were elevated in both solid and haematological malignancy patients, a further 15 CCGs were uniquely upregulated in solid tumours while only one CCG (IL-18) was uniquely upregulated in haematological malignancies. Also, only one CCG (angiopoietin receptor Tie-2) was also significantly downregulated (30-35%) in cancer patients and this was observed for both solid and haematological malignancy. Besides a function in angiogenesis, Tie-2 also controls cellular adhesion and invasion and therefore metastatic behaviour, and a downregulation of Tie-2 has been reported in squamous cell carcinomas in cell lines and tissue [34].

Alluding further on the differences between solid and haematological malignancies, we identified 3 CCGs that passed a stringent significance threshold for multiple comparison statistics for 55 CCGs. These 3 CCGs were SAA, VEGF-C and BDNF and were found to be higher in solid tumour patients. While a critical role of inflammation in progression of haematological malignancy including myeloid malignancies is now recognized [35], an increased acute phase marker protein SAA in solid tumours compared to haematological malignancy patients suggests that inflammation is more central to solid tumours. Similarly, an association of VEGF-C and BDNF with solid tumours is not surprising since tumour microenvironment remodelling with vascular growth aids in the growth and maintenance of solid tumours while being less pronounced in haematological malignancies [13,29,36].

In this background biology of CCG alterations in cancer patients, we further studied how SARS-CoV-2 exposure alters the CCGs in cancer patients. This was conducted as part of a prospective surveillance study covering the first and second COVID-19 wave in Belgium. Most of the studied patients were asymptomatic or had mild disease while only a minority required hospitalization. Keeping that in mind, we first describe a significant upregulation in levels of 17 CCGs in HCWs exposed to SARS-CoV-2 compared to unexposed HCWs while 3 CCGs were significantly downregulated (Tie-2, PlGF and IP-10). However, likely because CCGs were already highly elevated in the unexposed cancer groups, the effect on CCG alteration in SARS-CoV-2-exposed solid and haematological malignancy patients was less pronounced compared to the respective unexposed cancer groups. Only SAA and CRP were commonly elevated in exposed solid and haematological malignancies, and these represent the two most common acute phase protein markers employed in COVID-19 as well as in other inflammatory and infectious conditions and importantly suggest a prolonged pro-inflammatory state in exposed cancer patients [37]. Moreover, SAA has been previously shown to elicit immune evasion by the tumour through the reprogramming of macrophages to the pro-tumour M2 type and downregulation of anti-tumour responses [38].

In addition, we reported 2 CCGs (MCP-3 and IL-2) that were uniquely elevated and 5 CCGs (IL-9, VEGF-C, bFGF, BDNF and total TGF-β) that were uniquely downregulated in the solid tumour group exposed to SARS-CoV-2 compared to unexposed. While decreased VEGF-C, bFGF and BDNF could dysregulate angiogenesis in solid tumours, a decreased IL-9 and elevated MCP-3, SAA and IL-2 might help to accelerate progression of solid tumours. We also showed that IL-2 was amongst the selected cytokines that remained significantly elevated in the SARS-CoV-2 exposed solid tumour group over a 3-month study period. IL-2 is a major recruiter of T-helper cells, and also promotes self-tolerance by expansion of regulatory T cells [39]. Similarly, MCP-3 has been shown to help support the tumour microenvironment through the recruitment of tumour-associated lymphocytes and is also associated with infiltration of tumour-associated macrophages that aid immune evasion [40,41].

Conversely, patients with haematological malignancies showed a unique elevation for 5 CCGs (TNF-α, IFN-β, TSLP, soluble VCAM-1, and IP-10). While TNF-α, IFN-β, TSLP, SAA and sVCAM-1 are known to promote growth or enable immune evasion in haematological tumours [5,38,42-44], the unique elevation of IP-10 in haematological malignancies is also noteworthy as it promotes angiogenesis and metastasis in solid tumours and has also been found in increased levels in chronic myelomonocytic leukaemia patients [45,46]. Recent evidence has also emerged that highlights intriguing parallels between inflammatory pathways and aberrant immune cell crosstalk in metastasis formation and the role that primary tumours play in hijacking these interactions to enhance their metastatic potential [47]. Taken together, these data suggest that CCG profiles in haematological patients seem to be altered towards promotion of cancer progression after SARS-CoV-2 exposure. This is even more concerning as not only levels of CCGs such as MCP-3 and TNF-α show no decline till at least 3 months after SARS-CoV-2 exposure, levels of IL-2 and IP-10 even increase during this period. A recent study has also showed sustained immune dysregulation in haematological cancer patients displaying heterogeneous humoral responses and an exhausted T cell phenotype up to three months after in SARS-CoV-2 exposure [15]. Together these data prompt for an increased vigilance in clinical follow up for any sequelae of faster cancer progression after SARS-CoV-2 infection in patients with haematological malignancies, as has been speculated recently [48-50].

On the other hand, the question remains whether elevated baseline cytokine levels in cancer patients might offer protection in the initial stages of SARS-CoV-2 infection. While some studies have not reported increased COVID-19 severity in cancer patients [19], those that did have only considered hospitalized patients [16]. Many of the elevated cytokines in unexposed cancer patients in our study, especially with solid tumours, include pro-inflammatory Th1-related cytokines, which are classically targeted against bacteria and viruses. In our cancer cohort, SARS-CoV-2 exposure was only reported in approximately 4% of cancer patients compared to 3.1–6.9% in the overall Belgian population in the same time interval [51] and approximately 12% in the HCWs [26]. It is likely this can be explained by better self-protection of cancer patients. While cancer patients had more SARS-CoV-2 related mortality and severe/critical illness, 71% of the exposed solid cancer patients remained asymptomatic or mild to moderately ill, which was not significantly different from non-severe illness showed by HCWs (89.5%). This is noteworthy as HCWs in our study were, on average, 24 years younger and had fewer co-morbidities than cancer patients. It is tempting to speculate that high baseline levels of innate pro-inflammatory cytokines in cancer patients might be beneficial to rapidly deal with low-level exposure to SARS-CoV-2. However, it seems that once the balance of the “primed” immune system is disrupted, COVID-19 can be more severe and lethal in a subset of cancer patients.

As limitations, this study is a case-control study and prospective data collection especially for patients who were not positive for SARS-CoV-2 was not possible. Secondly, although older healthy controls were enrolled for this study, this group remained younger and had fewer co-morbidities compared to the cancer groups. And lastly, as the study was conducted in an oncology unit setting, cancer patients that were immediately transferred to the COVID-19 wards and had succumbed to the infection, were not included.

## Conclusions

We show here that cancer patients have intrinsically high levels of inflammatory cytokines/chemokines as well as angiogenic and other growth factors, that escalate significantly after SARS-CoV-2 infection, especially in haematological malignancy patients. Moreover, while cytokine profiles in patients with solid tumours stabilised over time, patients with haematological malignancies showed a sustained dysregulated immune response persisting for up to 3 months during the study period. As several of the cytokines/chemokines and growth factors studied here are also tumour promoting factors, our data calls for increased vigilance in patients with haematological malignancy with SARS-COV-2 infection as a part of long COVID-19 surveillance.

## Data Availability

All data produced in the present study are available upon reasonable request to the authors

## Supplementary Materials

Figure S1: Proportion of patients according to severity in solid and haematological malignancies as well as HCWs, Figure S2: Plasma levels of additional CCGs, Figure S3: Summarising figure showing cytokines significantly increased and decreased, Table S1: Patient characteristics, Table S2: Overview of all patients with information on cancer types, topography, morphology and behaviour of the malignancy, Table S3: Alterations in CCGs for unexposed solid and haematological malignancy patients compared to unexposed healthcare workers, Table S4: Alterations in CCGs levels in SARS-CoV-2-exposed individuals compared to unexposed individuals.

## Author Contributions

Conceptualization, S.K.-S. and P.V.D.; formal analysis, F.H.R.D.W., A.H., S.K.-S., E.R. and E.F.; investigation, F.H.R.D.W., A.H., A.K., B.’s.J., R.K.J., and V.V.a..; writing—original draft preparation, F.H.R.D.W., A.H. and S.K.-S..; writing—review and editing, all authors; funding acquisition, S.K.-S., M.P., and P.V.D. All authors have read and agreed to the published version of the manuscript.

## Institutional Review Board Statement

The study was conducted according to the guidelines of the Declaration of Helsinki and approved by the Ethics Committee of the Antwerp University Hospital (EC number 20/13/156, internal EDGE 001070).

## Informed Consent Statement

Informed consent was obtained from all subjects involved in the study.

## Data Availability Statement

The data that support the findings of this study are available from the corresponding author upon reasonable request.

