## Supplementary Materials for "Blood cytokine analysis suggests that SARS-CoV-2 infection results in a sustained tumour promoting environment in cancer patients"

**Supplementary Table 1: Patient characteristics***

|  | **Solid cancer (n = 68)** | | **Haematological cancer**  **(n = 38)** | | **HCW (n = 57)** | |
| --- | --- | --- | --- | --- | --- | --- |
|  | **n (%) or mean (SD)** | | **n (%) or mean (SD)** | | **n (%) or mean (SD)** | |
|  | **Exposed^a^** | **Unexp^b^** | **Exposed^a^** | **Unexp^b^** | **Exposed^a^** | **Unexp^b^** |
| Sleep apnoea | 3 (8.3%) | 1 (3.1%) | 1 (6.3%) | 0 (0%) | 0 (0%) | 1 (2.4%) |
| Cardiovascular disease | 3 (8.3%) | 6 (18.8%) | 2 (12.5%) | 3 (9.4%) | 1 (6.7%) | 0 (0%) |
| Thromboembolic disease | 2 (5.6%) | 3 (9.4%) | 2 (12.5%) | 1 (3.1%) | 0 (0%) | 0 (0%) |
| Renal disease | 1 (2.8%) | 1 (3.1%) | 1 (6.3%) | 0 (0%) | 0 (0%) | 0 (0%) |
| Pulmonary disease | 7 (19.4%) | 2 (6.3%) | 4 (25.0%) | 1 (3.1%) | 4 (26.7%) | 3 (7.1%) |
| Diabetes | 6 (16.7%) | 2 (6.3%) | 1 (6.3%) | 0 (0%) | 0 (0%) | 0 (0%) |
| Metabolic disease | 4 (11.1%) | 3 (9.4%) | 1 (6.3%) | 2 (6.3%) | 1 (6.7%) | 3 (7.1%) |
| Hypertension | 6 (16.7%) | 5 (15.6%) | 3 (18.8%) | 3 (9.4%) | 1 (6.7%) | 1 (2.4%) |
| Infection | 5 (13.9%) | 6 (18.8%) | 4 (25.0%) | 7 (21.9%) | 0 (0%) | 2 (4.8%) |
| Allergic constitution | 2 (5.6%) | 0 (0%) | 0 (0%) | 1 (3.1%) | 1 (6.7%) | 3 (7.1%) |
| Gastrointestinal disease | 2 (5.6%) | 6 (18.8%) | 4 (25.0%) | 3 (9.4%) | 0 (0%) | 4 (9.5%) |
| Autoimmune disorder | 3 (8.3%) | 0 (0%) | 1 (6.3%) | 1 (3.1%) | 1 (6.7%) | 2 (4.8%) |
| Recent non-COVID-19 vaccination | 0 (0%) | 1 (3.1%) | 0 (0%) | 0 (0%) | 0 (0%) | 0 (0%) |
| Epileptic & neurologic disorder | 1 (2.8%) | 1 (3.1%) | 1 (6.3%) | 1 (3.1%) | 1 (6.7%) | 0 (0%) |
| Bone disorders | 0 (0%) | 0 (0%) | 0 (0%) | 1 (3.1%) | 0 (0%) | 0 (0%) |
| * Consult Table 1 in main text for other patient variables  HCWs: Health care workers SD: standard deviation  ^a^All exposed individuals were selected  ^b^Unexposed individuals were group-matched to exposed individuals based on age, gender and cancer type in case of cancer patients | | | | | | |

**Supplementary Table 2:** Overview of all patients with information on cancer types, topography, morphology and behaviour of the malignancy. Last column represents the exposure to SARS-CoV-2 confirmed by either PCR or positive serology.

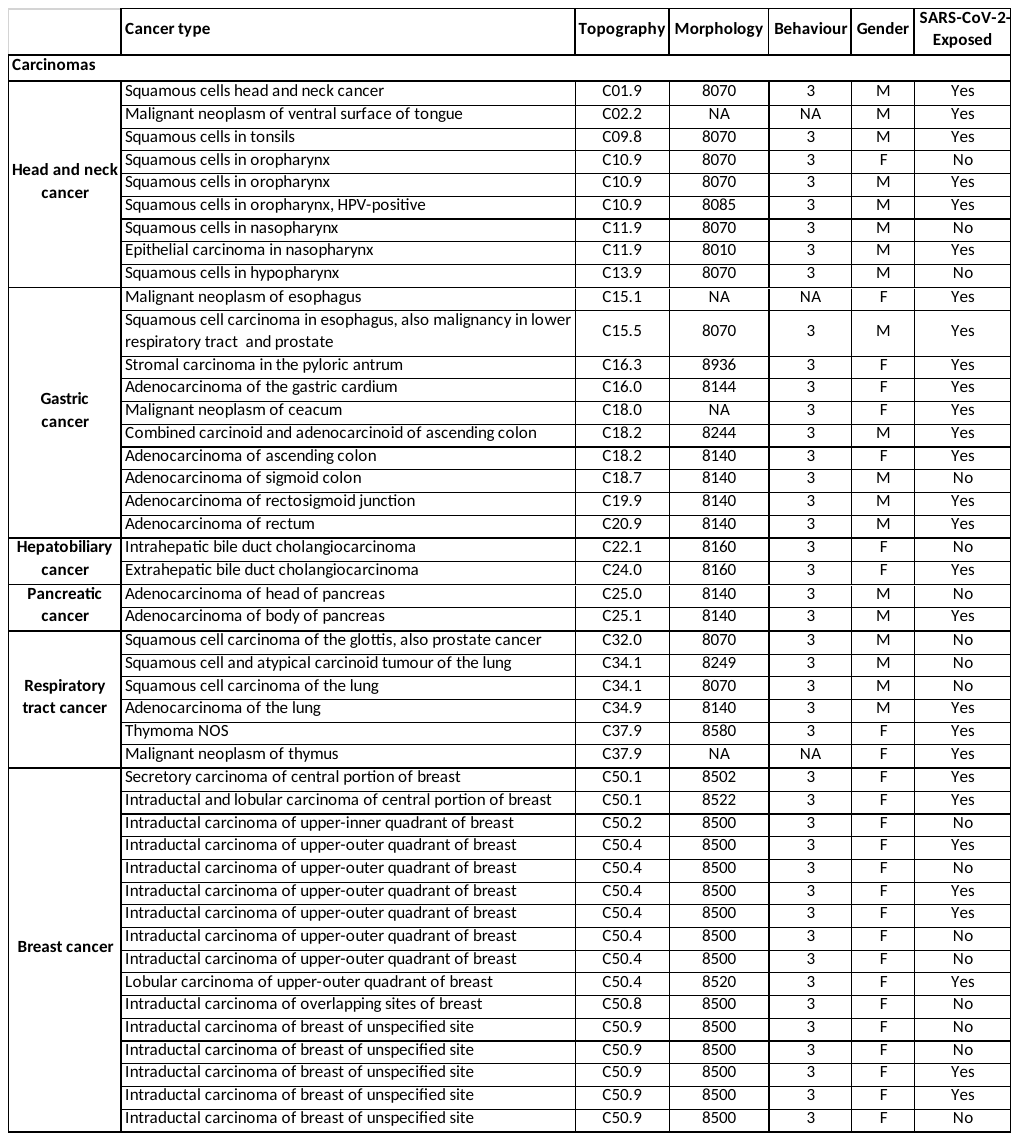

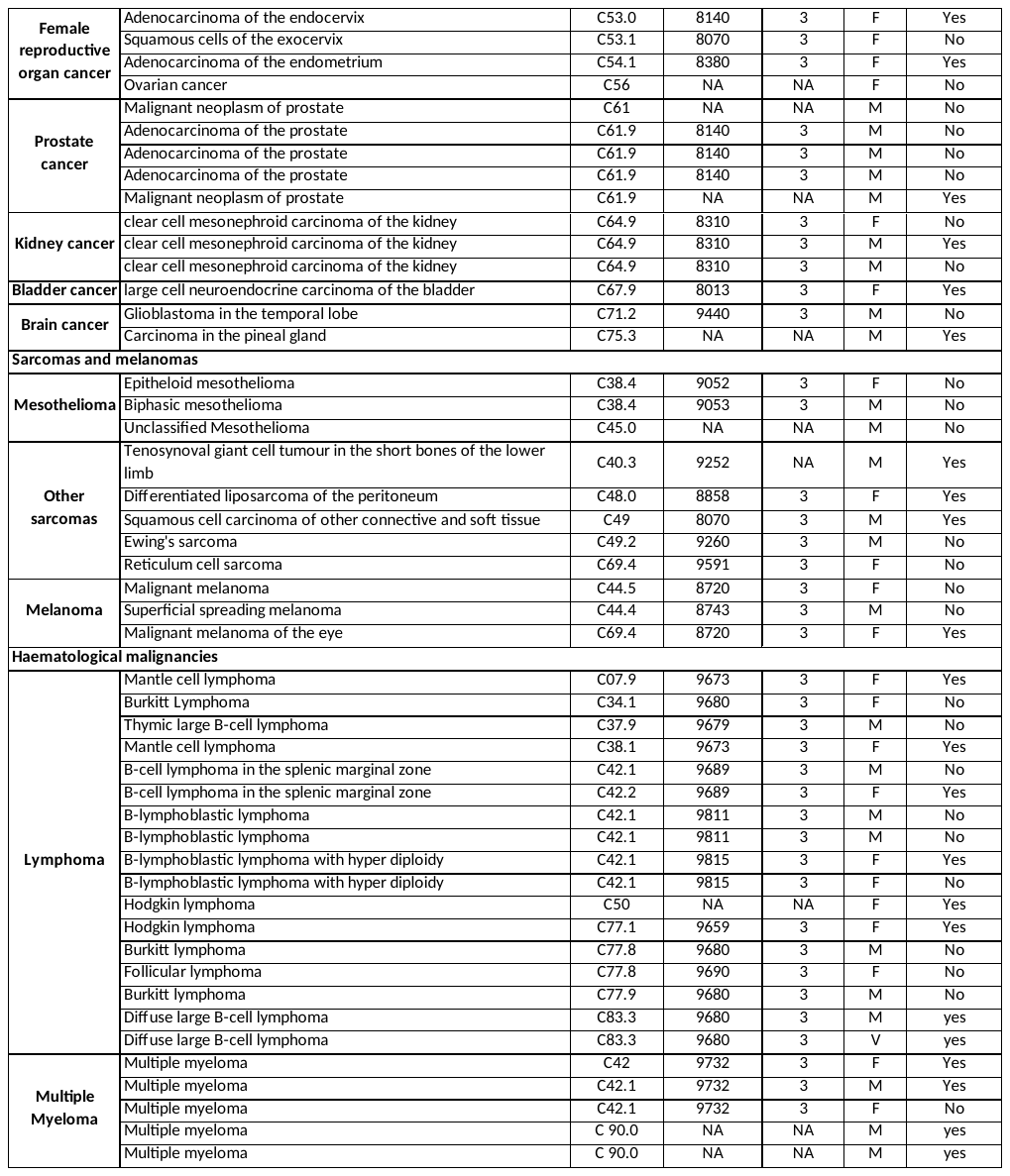

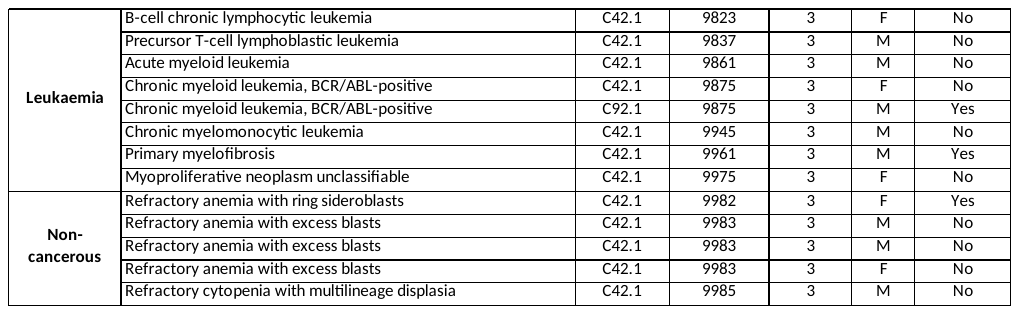

**Supplementary Table 3**: Alterations in CCGs for unexposed solid and haematological malignancy patients compared to unexposed healthcare workers (HCWs). The table represents the p-values and fold changes with one-way ANOVA between the three groups followed by pair wise comparisons with post-hoc Tukey correction for multiple comparisons.

|  | | **Solid cancer vs HCW** | | **Haematological cancer vs HCW** | |
| --- | --- | --- | --- | --- | --- |
| **CCG upregulated** | ***P* ANOVA** | **Fold Change Increase (95% CI)** | ***P* Tukey** | **Fold Change Increase (95% CI)** | ***P* Tukey** |
| **TNF-α** | < 0.001** | 1.3 (1.2 - 1.5) | < 0.001 | 1.3 (1.1-1.5) | 0.002 |
| **IL-6** | < 0.001** | 1.9 (1.6 - 2.3) | < 0.001 | 1.6 (1.3-2.0) | < 0.001 |
| **IL-1Ra** | 0.002** | 1.6 (1.2 - 2.1) | 0.003 | 1.5 (1.1-2.0) | 0.04 |
| **IL-8** | 0.008** | 1.8 (1.3 - 2.7) | 0.007 | 1.5 (0.9-2.2) | ns |
| **CRP** | 0.002** | 2.3 (1.5 - 3.6) | 0.002 | 1.3 (0.8-2.2) | ns |
| **SAA** | < 0.001** | 2.6 (1.6 - 4.1) | < 0.001 | 1.2 (0.7-2.0) | ns |
| **IFN-γ** | < 0.001** | 5.1 (3.3 - 7.9) | < 0.001 | 8.1 (5.0-13.2) | < 0.001 |
| **IL-18** | 0.029** | 1.3 (1.0-1.7) | ns | 1.5 (1.1-2.0) | 0.04 |
| **IL-17A** | < 0.001** | 1.8 (1.5 - 2.2) | < 0.001 | 1.7 (1.3-2.1) | < 0.001 |
| **IL-22** | < 0.001** | 1.2 (1.1 - 1.4) | 0.011 | 1.3 (1.1-1.5) | 0.001 |
| **IL-33** | 0.002** | 1.1 (1.0 - 1.2) | 0.003 | 1.1 (1.0-1.1) | 0.032 |
| **IL-5** | 0.006** | 1.4 (1.1 - 1.6) | 0.005 | 1.1 (0.9-1.4) | ns |
| **TSLP** | < 0.001** | 1.8 (1.5 - 2.1) | < 0.001 | 1.4 (1.1-1.7) | 0.011 |
| **IL-9** | < 0.001** | 1.4 (1.2 - 1.6) | < 0.001 | 1.2 (1.1-1.4) | 0.013 |
| **IL-21** | 0.017** | 1.4 (1.1 - 1.9) | 0.018 | 1.3 (1.0-1.7) | ns |
| **Eotaxin** | 0.043 | 1.3 (1.1 - 1.7) | 0.037 | 1.1 (0.9-1.4) | ns |
| **IL-10** | < 0.001** | 1.3 (1.2 - 1.4) | < 0.001 | 1.2 (1.1-1.4) | < 0.001 |
| **IL-2Rα** | 0.002** | 1.5 (1.2-1.9) | 0.002 | 1.3 (1.0-1.7) | ns |
| **IL-15** | 0.004** | 1.2 (1.0-1.4) | 0.032 | 1.3 (1.1-1.5) | 0.009 |
| **VEGF-A** | 0.039 | 1.8 (1.2-3.0) | 0.034 | 1.3 (0.7-2.1) | ns |
| **bFGF** | 0.056 | 2.7 (1.2-6.0) | 0.048 | 1.4 (0.6-3.5) | ns |
| **PlGF** | < 0.001** | 1.4 (1.2-1.7) | 0.001 | 1.5 (1.2-1.8) | 0.002 |
| **Flt-1** | 0.027** | 1.3 (1.1-1.6) | 0.031 | 1.2 (1.0-1.5) | ns |
| **EPO** | < 0.001** | 2.1 (1.5-2.8) | < 0.001 | 2.3 (1.6-3.2) | < 0.001 |
| **sICAM-1** | 0.017** | 1.1 (1.0-1.1) | 0.031 | 1.1 (1.0-1.1) | ns |
| **G-CSF** | < 0.001** | 1.8 (1.4-2.4) | < 0.001 | 1.7 (1.2-2.2) | 0.003 |
| **M-CSF** | < 0.001** | 1.5 (1.2-1.8) | 0.001 | 1.3 (1.1-1.7) | 0.033 |
| **GM-CSF** | 0.015** | 1.1 (1.0-1.1) | 0.015 | 1.0 (1.0-1.1) | ns |
| **MCP-1** | < 0.001** | 1.7 (1.4-2.0) | < 0.001 | 1.6 (1.4-1.9) | < 0.001 |
| **MCP-2** | 0.003** | 1.3 (1.1-1.6) | 0.024 | 1.4 (1.1-1.8) | 0.007 |
| **MCP-3** | 0.012** | 1.3 (1.1-1.6) | 0.01 | 1.1 (0.9-1.3) | ns |
| **MIP-1β** | 0.035 | 1.4 (1.1-1.9) | 0.036 | 1.3 (0.9-1.7) | ns |
| **MIP-3α** | 0.013** | 1.5 (1.1-1.9) | 0.012 | 1.1 (0.8-1.5) | ns |
| **Fractalkine** | < 0.001** | 1.3 (1.1-1.5) | 0.004 | 1.3 (1.1-1.5) | 0.003 |
| **CTACK** | 0.004** | 1.29 (1.1-1.5) | 0.009 | 1.27 (1.1-1.5) | 0.033 |
| **CCG downregulated** | ***P* ANOVA** | **Fold Change Decrease (95% CI)** | ***P* Tukey** | **Fold Change Decrease (95% CI)** | ***P* Tukey** |
| **Tie-2** | <0.001** | 1.4 (1.2-1.5) | <0.001 | 1.3 (1.1-1.5) | 0.001 |
| HCW: Health care workers  **FDR-corrected significance (q-value) <0.05 | | | | | |

**Supplementary Table 4: Alterations in CCGs levels in SARS-CoV-2-exposed individuals compared to unexposed individuals.** A) healthcare workers (HCWs), B) solid cancer patients and C) haematological malignancy patients. T-test was performed on Log10 transformed data.

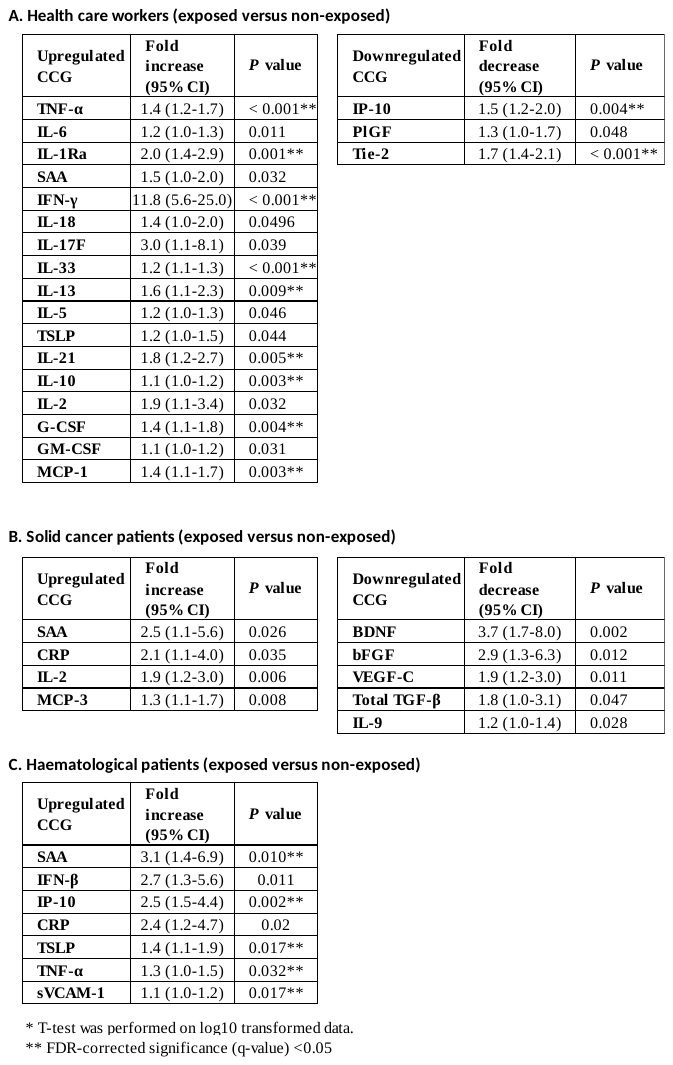

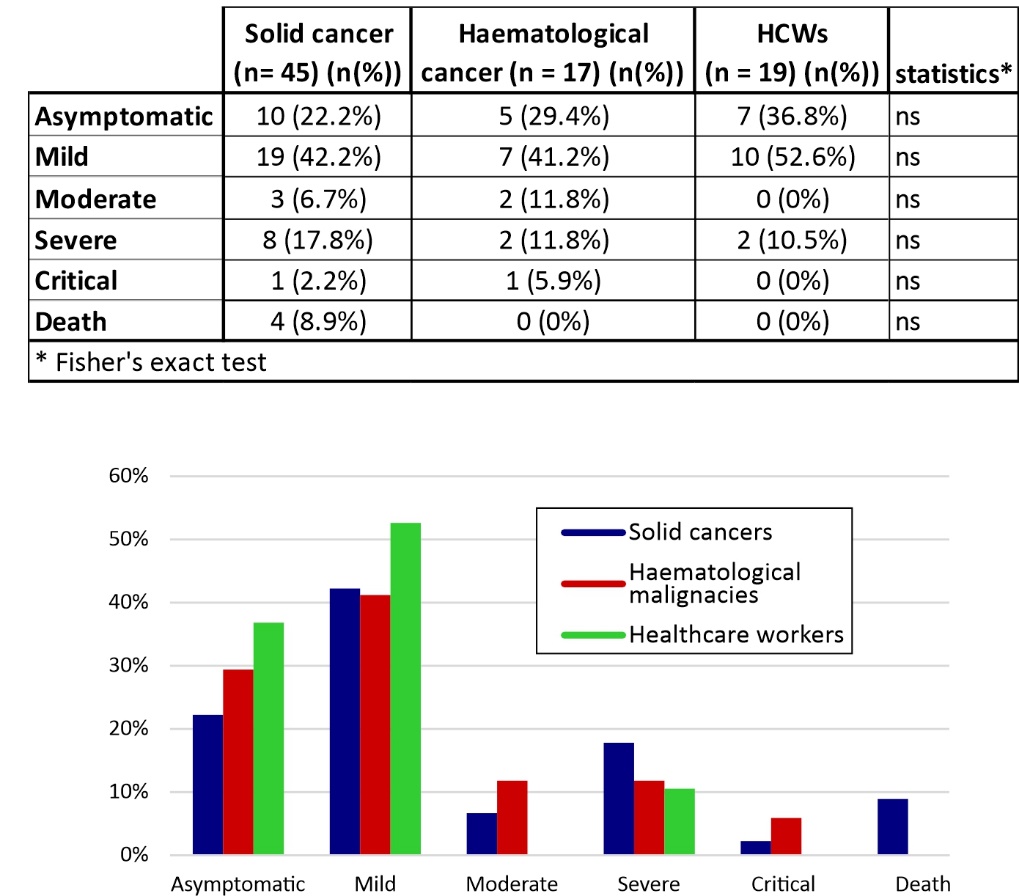

**Supplementary Figure 1:** Proportion of patients according to severity in solid and haematological malignancies as well as HCWs. The upper panel shows the precise numbers and the statistics.

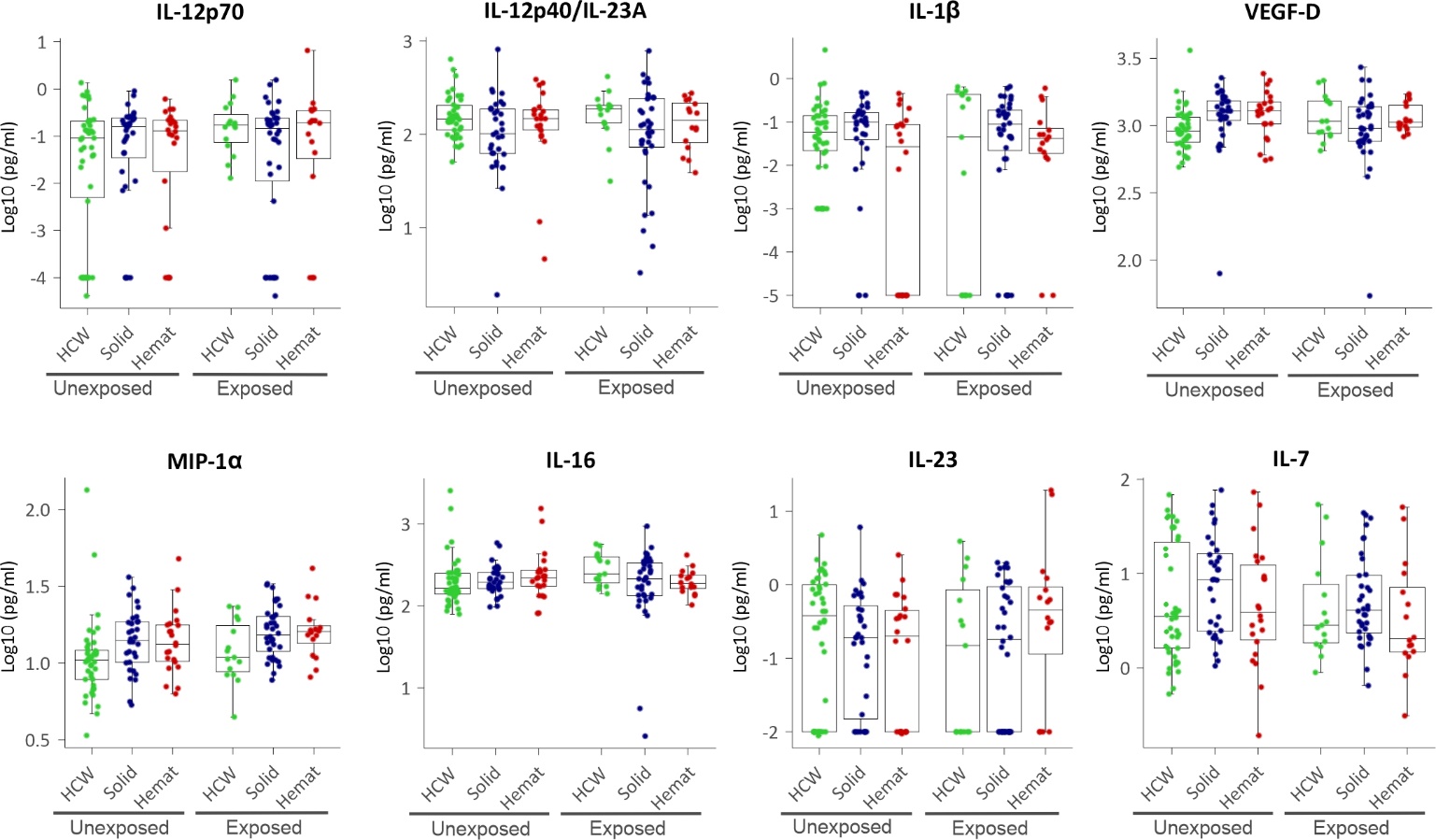

**Supplementary Figure 2.** **Plasma levels of additional CCGs.** HCW, health care workers (green), Solid, patients with solid tumours (blue), Hemat, patients with haematological malignancies (red). Each dot represents the sample closest to exposure in case of exposed patients, and an average of multiple timepoints, when available, for unexposed patients.

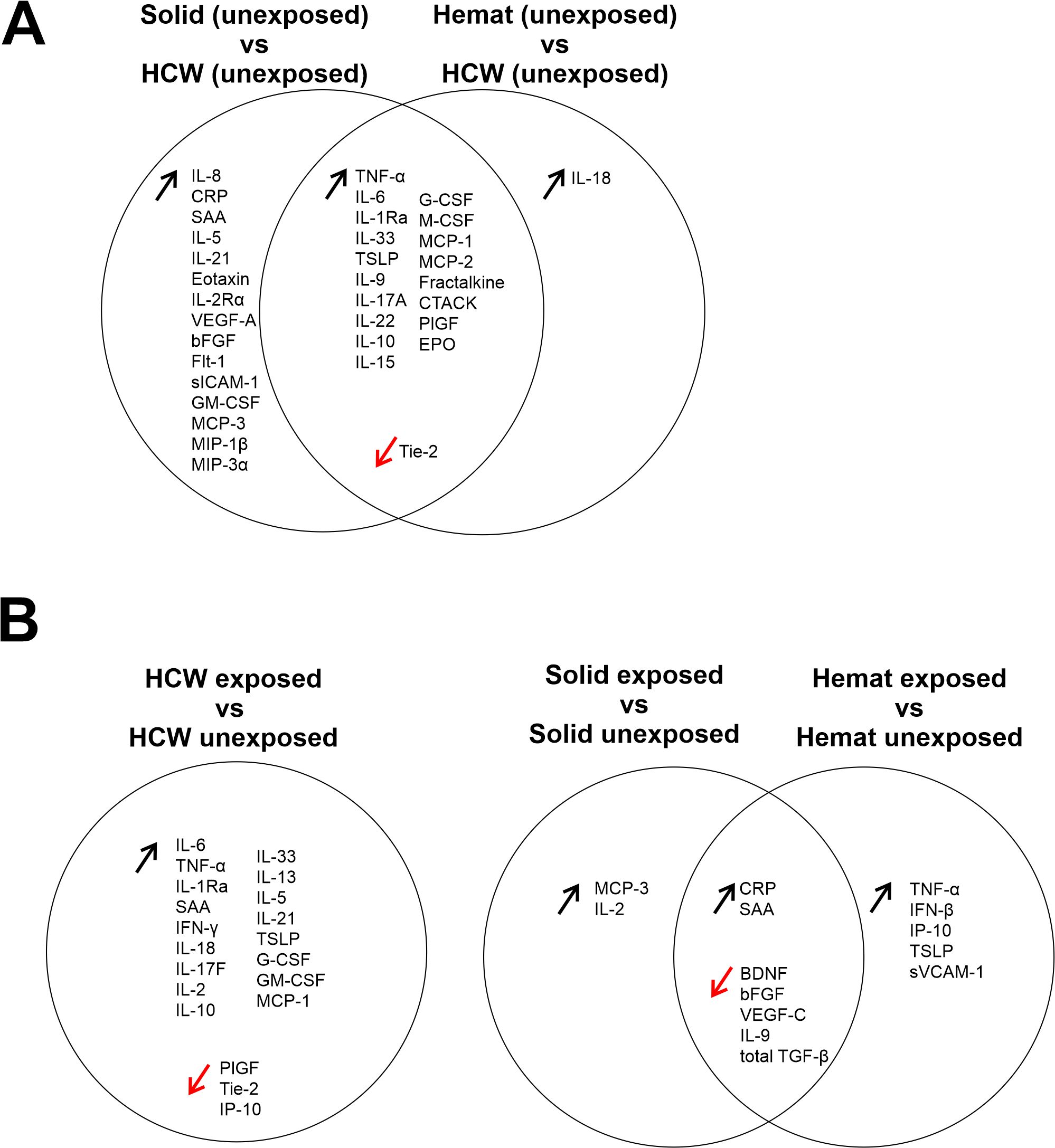

**Supplementary Figure 3:** Summarising figure showing cytokines significantly increased and decreased **A)** in relatively unexposed patients with solid cancers and with haematological malignancies compared to unexposed HCWs. **B)** Cytokines significantly increased and decreased in HCWs exposed to SARS-CoV-2 compared to unexposed HCWs and increased and decreased in SARS-CoV-2 exposed solid and haematological malignancy patients compared to unexposed solid and haematological malignancy patients. HCW, health care workers, Solid, patients with solid tumours, Hemat, patients with haematological malignancies.
